# Perceived Satisfaction in Fetal Therapy: Proposal and Pilot of a Novel Assessment Tool (*FETAL Surgery Global Satisfaction*)

**DOI:** 10.1101/2025.08.11.25333341

**Authors:** Marta Domínguez-Moreno, Ángel Chimenea, José De-Martín-Hernández, Lutgardo García-Díaz, Guillermo Antiñolo

## Abstract

**Background:** The quality of healthcare services has garnered significant attention within the medical field, with patient-perceived satisfaction emerging as a crucial indicator. Analyzing patient satisfaction could uncover areas of weakness, suggesting the need for improvements in care. However, in the specialized field of Fetal Medicine and Therapy, there lacks a validated tool for assessing care quality from the patient’s perspective. This study aims to design and pilot an instrument for measuring patient-perceived satisfaction as a barometer of the quality of services provided.

**Methods:** We developed a survey instrument to assess six key areas of healthcare (Follow-up care, environment, transparency, accessibility of care, link between physician and patient, and global satisfaction), through 20 multiple-choice questions. It was initially piloted on a sample of 34 pregnant women undergoing a specific fetal therapy (Ex Utero Intrapartum Treatment, EXIT) at our center between June 2007 and January 2024.

**Results:** A total of 29 out of 34 patients agreed to participate. According to the proposed tool and the suggested scoring scale, we achieved an overall score of 4.67, indicating a very satisfactory rating. The highest mean score was for the “Global satisfaction” domain (4.76), revealing that participants where highly likely to recommend our department. It was followed by “Follow-up care” and “Environment” domains (4.69, each). Notably, “Transparency” and “Link between physician-patient” were the least rating domains (4.58 and 4.62, respectively), suggesting the timely provision of enhancement.

**Conclusions:** We propose “*FETAL Surgery Global Satisfaction Tool*” as a potential valuable instrument to assess patient-perceived satisfaction. It could provide insights into the quality of services offered by Fetal Medicine and Therapy Units. By identifying key areas for improvement, this tool could support continuous quality enhancement across Fetal Therapy programs globally. Further studies are warranted to evaluate its reliability and validity.

## 1. Background

In recent years, there has been a growing interest in the medical field regarding the quality of provided medical services^1^. Simultaneously, healthcare centers implement strategies related to the organization and design of medical care processes, with particular emphasis on covering patients ’needs and expectations.

Patient-perceived satisfaction is now widely regarded as a crucial indicator of medical care quality^2–4^ and must be a priority in any healthcare system committed to continuous improvement and excellence^5^. It is defined as the difference between an individual’s perceived and an expected performance^1^. By gathering information on patients’ perceptions of the care received, it becomes possible to determine the level of satisfaction or dissatisfaction in relation to their expectations from the organization and their experiences with it^2^.

The Fetal Medicine and Therapy Units are recognized as pivotal services of tertiary-level maternal-fetal medicine hospitals. All complex fetal pathologies are referred to them, for precise antenatal diagnosis and accurate management options. In this context, parents face a range of uncertainties regarding the future of their unborn child and the potential maternal risks, so the options provided pose major ethical challenges^6^.

Given the worldwide and yearly increase in the number of centers specializing in Fetal Medicine and Therapy^7^, enabled by advancements in imaging, surgical instrumentation, and techniques^8,9^, the demands and expectations for its convenience are also growing. Thus, it becomes necessary to analyze patient-perceived satisfaction as a reflection of the quality of the services provided. However, a specific evaluation of patients’ satisfaction in that field has not been previously published.

Accordingly, we sought to determine perceived satisfaction among patients enrolled in a Fetal Therapy program, by designing and pilot a novel measuring tool. Specifically, we propose a structured survey as a barometer of the care quality. Through its implementation, it would be feasible to identify areas for improvement and introduce strategies seeking to enhance them.

## 2. Methods

### 2.1. Study-design steps

We designed a survey instrument to analyze satisfaction perceived by patients undergoing Fetal Therapy on account of fetal pathologies.

#### Literature review

A comprehensive and extensive review of the literature was conducted to identify key areas influencing patient-perceived satisfaction in the context of Fetal Medicine. A search for validated instruments on this field in PubMed and Google Scholar yielded no results. However, we analyzed relevant studies and existing validated tools focused on overall patients satisfaction^1,3,10–17^, to select survey domains. They would then need to be adapted to fit the context of the Fetal Medicine and Therapy.

#### Focus groups and interviews

Focus groups and interviews were conducted with experts in Fetal Medicine, patients, and healthcare professionals to ensure a diverse range of perspectives. These rounds aimed to gather insights, discuss priorities, and achieve consensus on the items of interest to include in the survey. Participants included 10 healthcare professionals (including obstetricians, pediatric surgeons, and neonatologists), five patients who had previously undergone fetal surgery, and five members of the hospitaĺs quality improvement team.

#### Question development

Based on the literature review and insights from focus groups, 38 potential survey items were initially identified, categorized into six domains.

An iterative process of refinement was employed, achieving consensus among experts through the Delphi method, in accordance with the Conducting and Reporting Delphi Studies (CREDES) checklist^18^. It involved multiple rounds of questioning where experts rated and commented on each item until a high level of agreement was reached. Hence, the domains and questions of interest were prioritized. This process resulted in the removal of 18 questions, reaching to a final set of 20 questions for further review. Specific criteria for inclusion and exclusion of questions were based on relevance, clarity, and importance as rated by experts.

#### Pilot testing

The survey was pilot-distributed to evaluate clarity, identifying, and clarifying any awkward terms or ambiguous questions, and ensure it was understandable and covered all relevant aspects of patient-perceived satisfaction. Feedback was collected through follow-up interviews and surveys, and some minor necessary adjustments were made. This adaptation customized the questionnaire for investigating the perceived satisfaction among patients enrolled in a Fetal Therapy program.

We used a purposive sampling approach to achieve an adequate representation from patients who received a specific Fetal Therapy. The sample consisted of all pregnant women followed-up for a fetal anomaly by the Department of Maternal-Fetal Medicine, Genetics, and Reproduction of the Virgen del Rocío University Hospital, who underwent “Ex - Utero Intrapartum Treatment” (EXIT), from June 2007 to January 2024.

The respondents were not financially compensated. Those who did not complete or submit the survey were all excluded.

### 2.2. Proposed instrument

By following the steps outlined above, we proposed that patients-perceived satisfaction could be categorized into six domains:

1. **F**ollow-up care (3 questions)
2. **E**nvironment (5 questions)
3. **T**ransparency (3 questions)
4. **A**ccessibility of care (3 questions)
5. **L**ink between physician and patient (4 questions)
6. **Global satisfaction** (2 questions)

These domains were evaluated through 20 multiple-choice questions. The survey format included a general information section briefly describing the study’s purpose, followed by six blocks of questions, corresponding to the identified domains. Additionally, we included a free text-section at the end of the questionnaire looking at what the respondent liked about the Unit, what they felt needed improving or any other comments.

We proposed naming this new evaluation tool “*FETAL Surgery Global Satisfaction*”, based on the initials of each evaluated domain. The complete survey with each drafted question is shown in *Table 1*.

**Table 1.** FETAL Surgery Global Satisfaction Tool.

|  |  |  |  |  |  |
| --- | --- | --- | --- | --- | --- |
| **A1. Follow-up care** |  |  |  |  |  |
| Q1. How would you rate the frequency with which you were evaluated by the same professional of the Unit in successive medical visits? | 1 | 2 | 3 | 4 | 5 |
| Q2. How would you rate the participation of other specialists (pediatric surgeons, neonatologists...) as part of the multidisciplinary management provided by the Unit? | 1 | 2 | 3 | 4 | 5 |
| Q3. To what extent do you consider that the information provided by the different health professionals was consistent and congruent? | 1 | 2 | 3 | 4 | 5 |
| **A2. Environment** |  |  |  |  |  |
| Q4. How would you rate the condition of the Unit's facilities (cleanliness, tidiness, appearance, comfort)? | 1 | 2 | 3 | 4 | 5 |
| Q5. The waiting time to be attended by the professionals of the Unit was... | 1 | 2 | 3 | 4 | 5 |
| Q6. To what extent did you find the environment of the consultation pleasant, quiet and uninterrupted? | 1 | 2 | 3 | 4 | 5 |
| Q7. How would you rate the care of the Unit's medical and nursing staff with your privacy and confidentiality during the medical visit? | 1 | 2 | 3 | 4 | 5 |
| Q8. To what extent do you consider the appearance of the Unit's personnel and the care of their image to be adequate? | 1 | 2 | 3 | 4 | 5 |
| **A3. Transparency** |  |  |  |  |  |
| Q9. To what extent do you consider that you received clear and understandable information about your baby's suspected pathology and its treatment from the professionals of the Unit? | 1 | 2 | 3 | 4 | 5 |
| Q10. To what extent were you able to verify that the professionals of the Unit had adequate means for the understanding of the information given, including the preparation of drawings and explanatory diagrams? | 1 | 2 | 3 | 4 | 5 |
| Q11. How would you rate the frequency with which the Unit professional asked you questions to make sure that you understood the information given? | 1 | 2 | 3 | 4 | 5 |
| **A4. Accessibility of care** |  |  |  |  |  |
| Q12. After the suspicion of a pathology in the baby that required a more detailed study, do you consider that the referral to the Fetal Medicine and Therapy Unit was adequate in time and form? | 1 | 2 | 3 | 4 | 5 |
| Q13. In case you had a problem with the date or time of the assigned appointment, how would you rate the willingness of the Unit's staff to provide you with a new appointment? | 1 | 2 | 3 | 4 | 5 |
| Q14. Were the directions and signage available in the hospital appropriate for you to find your way around and get to the Unit? | 1 | 2 | 3 | 4 | 5 |
| **A5. Link between physician and patient** |  |  |  |  |  |
| Q15. How would you rate the kindness and courtesy of the professionals of the Unit who attended you? | 1 | 2 | 3 | 4 | 5 |
| Q16. How would you rate the trust, honesty and security that the professionals of the Unit conveyed to you? | 1 | 2 | 3 | 4 | 5 |
| Q17. To what extent did the professionals at the Unit show interest in knowing your emotions and understanding your needs? | 1 | 2 | 3 | 4 | 5 |
| Q18. As a patient, to what extent did you feel supported and accompanied by the medical team? | 1 | 2 | 3 | 4 | 5 |
| **A6. Global satisfaction** |  |  |  |  |  |
| Q19. Indicate your degree of satisfaction, in general terms, with the Fetal Medicine and Therapy Unit and the professionals who integrate it. | 1 | 2 | 3 | 4 | 5 |
| Q20. Based on your entire experience in the Fetal Medicine and Therapy Unit of our center, to what extent would you recommend it to pregnant women in a similar situation to yours? | 1 | 2 | 3 | 4 | 5 |
| Q21. Do you identify any aspect that could be improved? Point out any aspect of the Unit or of the professionals who compose it that, in your opinion, could be improved. Explain briefly. |  |  |  |  |  |

### 2.3. Scoring scale

A five-point Likert scale, known for its symmetry, equidistance, and inclusion of a neutral response option, was used for each question, with 5 indicating “very satisfactory” and 1 indicating “very unsatisfactory” rate. The proposed scale is as follows:

1. Very unsatisfactory
2. Unsatisfactory
3. Neutral, neither satisfactory nor unsatisfactory
4. Satisfactory
5. Very satisfactory

As not all domains included the same number of questions, we proposed calculating the score for each one as the average of the questions within it (*Table 2*). Every domain would consequently be assigned a score ranging from 1 to 5.

**Table 2.**
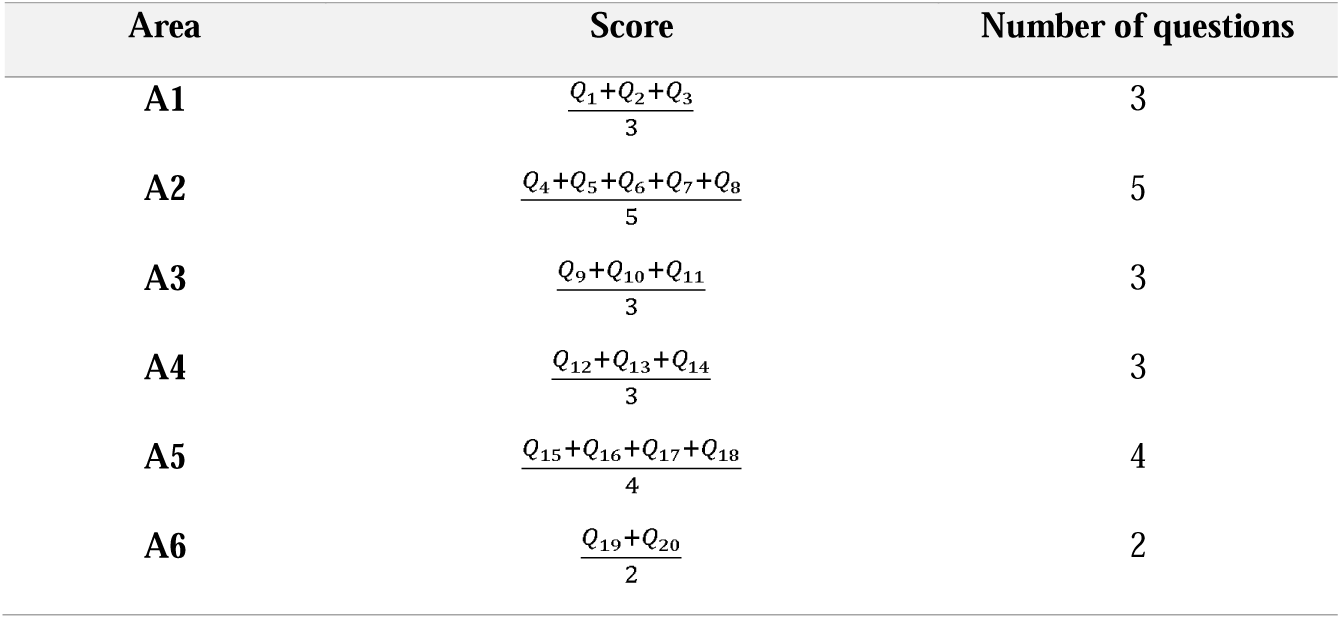
Scoring scale proposed.

Overall score would be calculated by dividing the total score by the number of domains (six), resulting in scores from 1 to 5. As all questions were written as positive statements, higher scoring represented a greater patient-perceived satisfaction. To facilitate the analysis, we proposed grouping scores into five major categories based on ranges. They are detailed in *Figure 1*.

**Figure 1.**
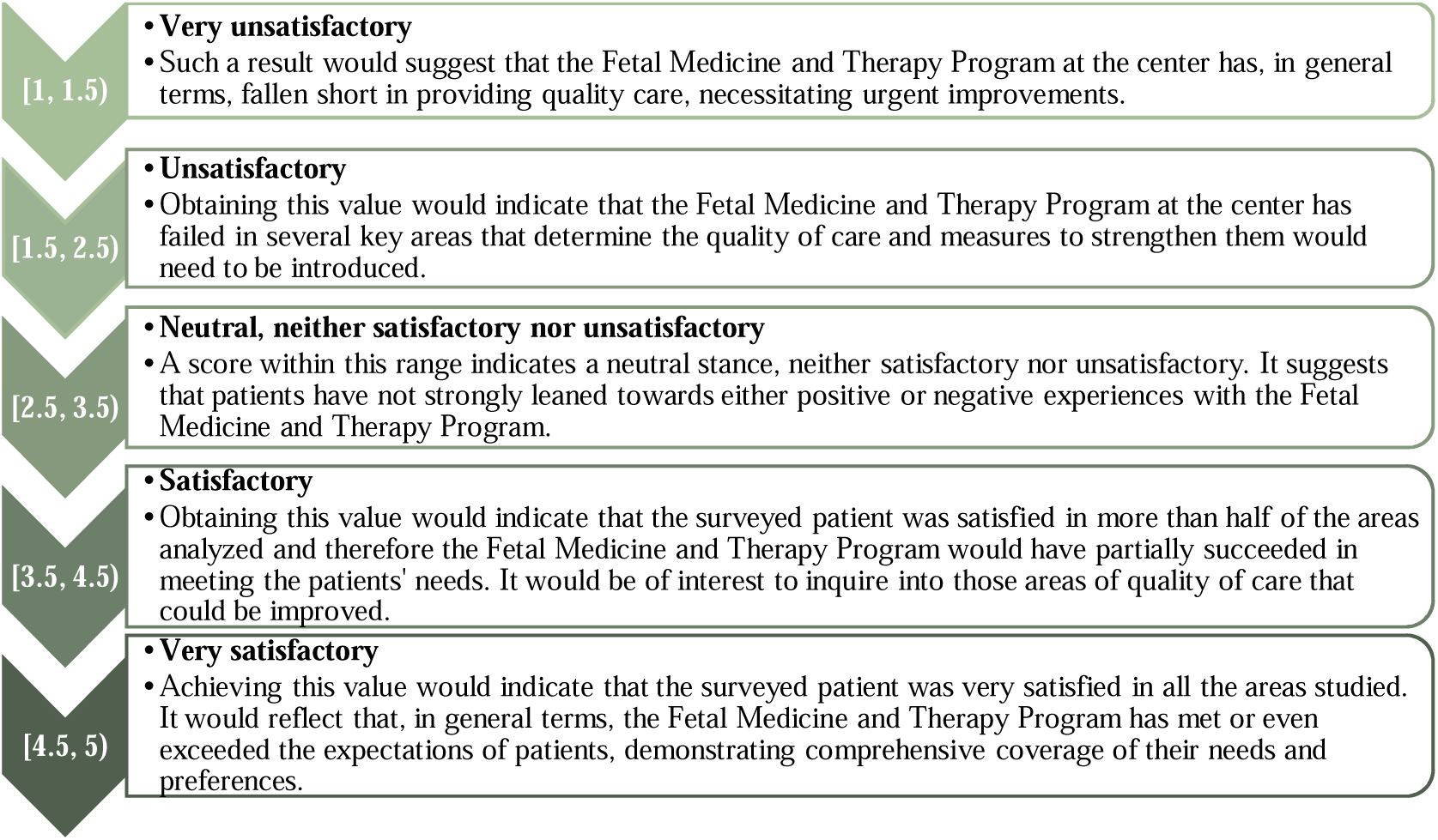
Satisfaction categories according to scores.

### 2.4. Tool administration procedure

The survey was self-administered, open-ended, and confidential. Following an initial telephone call explaining the survey’s objective and how to participate, it was uploaded online using Google Forms and distributed electronically via social media phone application to consenting women.

Participation was voluntary and informed consent was obtained. Patients provided identification when completing the survey, which was then anonymized to mitigate bias. Its completion was estimated to take approximately 5-7 minutes. Respondents were encouraged to carefully read the survey and received contact information from the healthcare provider for assistance with any queries. They were also offered to receive the survey results, which would be later analyzed and published.

To increase participation, a gentle reminder was sent out to non-responders with 2-weeks interval between the first contact.

Data collection took place in a designated area within our Department to ensure the confidentiality of the information obtained. It was also collected clinical and social data about patients: age, BMI, medical history, neonatal outcomes, and potential complications, obtained from the digital health record.

### 2.5. Data processing and analysis

Descriptive statistical analysis was performed, yielding specific results for each response/category. Variables were summarized using frequencies and proportions. Data was presented graphically to identify potential areas for improvement.

### 2.6. Ethical considerations

Prior to initiating data collection, we sought guidance from the institutional ethics committee (Andalusian Ethics Committee), which confirmed that formal authorization was unnecessary. Approval was obtained from the Quality Department of our institution, along with written institutional permission. Rigorous adherence to the principles outlined in the Declaration of Helsinki was maintained throughout every phase of the study. Written consent was obtained from each participant after introducing the questionnaires and receiving their consent.

## 3. Results

### 3.1. Demographic information

29 out of 34 women who underwent EXIT at our department completed the survey. The missing data rate was 14.7%, two patients declining to participate, and three others were not reachable with the available contact information.

Demographic characteristics are presented in *Table 3.* The median maternal age was 31.5 years. Of the total pregnant women undergoing EXIT, 17 were primigravida, and 5 had a previous caesarean section. Congenital diaphragmatic hernia was the most common indication (n=19). The median gestational age at the time of EXIT was 37+5 weeks of gestation. The median maternal and fetal operation time were 60 and 8.5 minutes, respectively. Following EXIT, none of the women suffered from surgical complications such as endometritis, puerperal fever, placental abruption, uterine dehiscence, or rupture.

**Table 3.** Demographic characteristics.

| Variables | EXIT (n=34) |
| --- | --- |
| Maternal age (years), median (range) | 31.5 (18-43) |
| Gravity, median (range) | 2 (1-7) |
| Parity, median (range) | 0.5 (0-6) |
| Previous C-section, n (%) | 5 (14.71) |
| GA at delivery (days), median (range) | 263 (216-273) |
| Maternal operation time (minutes), median (range) | 60 (35-180) |
| Fetal operation time (minutes), median (range) | 8.5 (3-24) |

### 3.2. Survey results

Our pilot analysis revealed an overall score of 4.67, indicating a very satisfactory rating.

The “global satisfaction” domain scored the highest (4.76). Surveyed participants were asked about their overall satisfaction with the Fetal Medicine and Therapy Program (mean 4.72) and, from a practical standpoint, whether they would recommend it to pregnant women in a similar situation to their own (mean 4.79).

The “follow-up care” and “environment” domains also scored highly (4.69 each). However, “Transparency” and “link between physician and patient” domains, while still positive, were identified as areas for potential improvement, scoring 4.58 and 4.62, respectively.

Individual score of the six evaluated domains is presented in *Table 4*.

**Table 4.** Individual score of evaluated areas.

| Areas | Score |
| --- | --- |
| A1. Follow-up care | 4.69 |
| A2. Environment | 4.69 |
| A3. Transparency | 4.58 |
| A4. Accesibility of care | 4.67 |
| A5. Link between physician-patient | 4.62 |
| A6. Global satisfaction | 4.76 |

Each question score with its mean and standard deviation is detailed in *Figure 2*.

**Figure 2.**
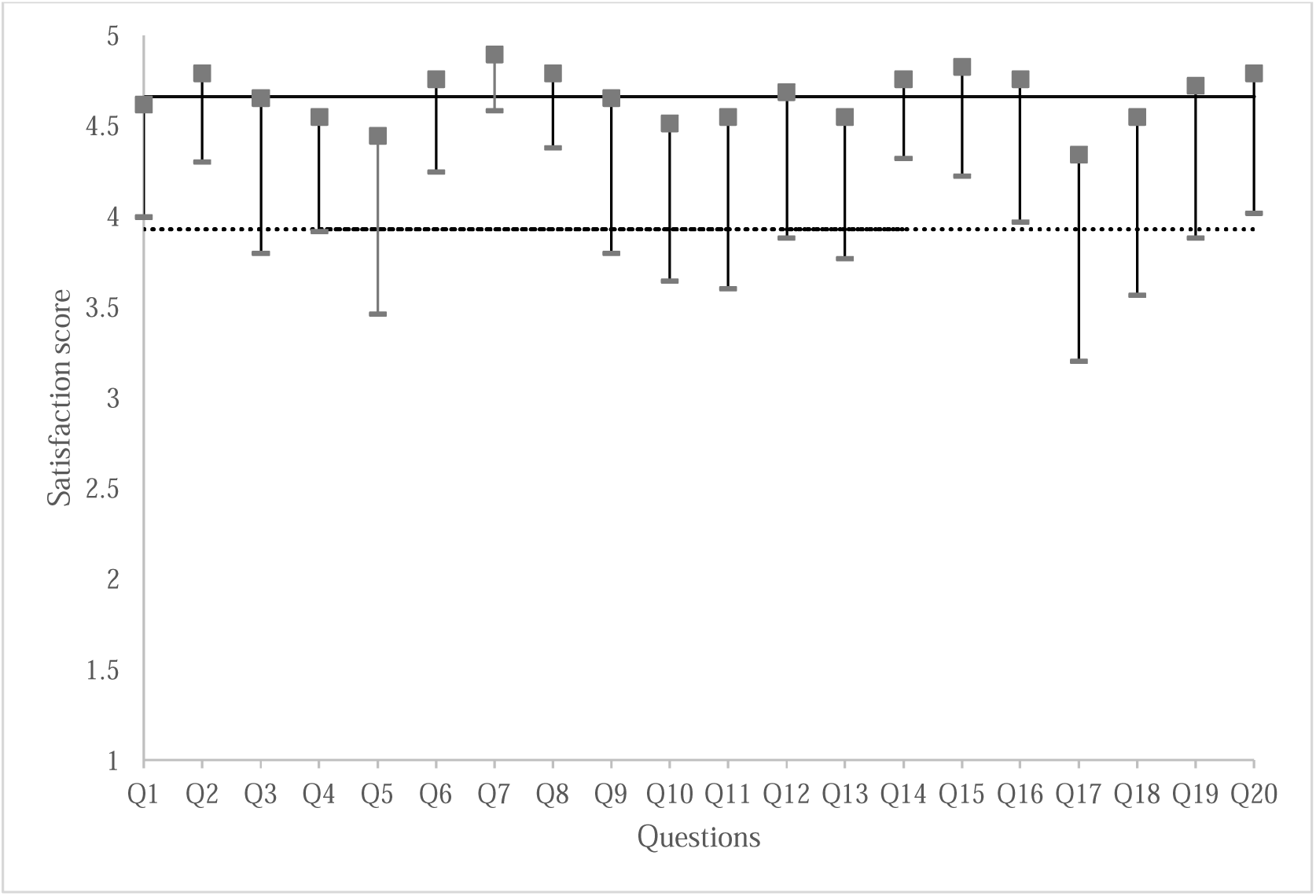
Satisfaction score for each question. Mean satisfaction score were calculated for each question based on all answers obtained and is represented on the ordinate axis. In the abscissa axis, it is shown each question, numbered from 1 to 20 (Q1 to Q20). The mean value of each response is represented by a square and the standard deviations are framed as two horizontal stripes extending from the ends of each square. The horizontal line represents the mean of all answers. The average of the “mean - standard deviation” of all questions is represented by a horizontal line composed of dots.

## 4. Discussion

### 4.1. Principal findings

This study provides an initial exploration of patient-perceived satisfaction within a Fetal Medicine and Therapy program using a novel assessment tool.

The high overall satisfaction score suggest that most patients felt positively about their care, particularly in terms of “environment” and “follow-up care”. However, the slightly lower score in “transparency” and “link between physician and patient” domains highlight the opportunities for further enhancement in certain areas, such as communication skills and emotional support.

To the best of our knowledge, this is the first study focused on perceived satisfaction among pregnant women undergoing Fetal Therapy.

### 4.2. Quality of care

Defined as “*the degree to which health services for individuals and populations increase the likelihood of desired health outcomes and are consistent with current professional knowledge*”^19^, it is a continuum from minimally acceptable to optimal care^19^.

In recent years, given the trend towards place the patient at the heart of care evaluation, there has been a growing interest in its measurement, stemming from different perspectives^20^. On one front, consumers want to be sure that they are cared for by doctors that will maximally help them^19^. Patients have higher expectations of the healthcare system, are better informed and access to more information^21^.

On the other side, healthcare providers aspire to ensure that their care is of a high level^19^. Then, to be aware of the quality care, it is crucial to measure it. In its absence, it would not be possible to identify areas for improvement^19^.

For decades, the assessment of care quality has focused on easily measurable and objective parameters from healthcare databases and safety guidelines, such as survival or mortality rates^20,22^. However, these instruments lack in fully capturing the subjective patient experience. In fact, they often fall short in addressing essential factors like hospital environment, continuity of care and emotional support^23^.

In this scenario, there is a need for tools to assess patients ’experience of the services provided. They can be used to benchmark the care quality and therefore, to suggest modifications or further developments^24^.

### 4.3. Patient satisfaction

Satisfaction is a multi-dimensional concept concerned with individual subjective perception of the quality and efficacy of services^24^. It is widely recognized as a pivotal component of care quality within healthcare organizations, offering a quantifiable metric for evaluation^1,2,25^.

Recent surveys in the UK have revealed a concerning trend: a record-low satisfaction level with healthcare services. Notably, the National Health Service (NHS) reported an overall satisfaction stood at a mere 29% in 2022^5^. Concerned about this situation, health systems try to introduce measures for change.

Unsurprisingly, the field of Fetal Medicine and Therapy faces the same challenges. Robust data on quality care and satisfaction in this area are lacking so far. Furthermore, there is currently no validating objective grading method to analyze patient-perceived satisfaction in this field.

The need for sensitive-context tool in this area derives from the significantly difference that could be observed between patients followed in these Units and other general patients. These cases are less common and more complex than in other specialties, involve an intense and close physician-patient interaction during the whole pregnancy, and above all, Fetal Medicine is an emerging area with a lack of collecting and analysis long-standing data. Thus, its evaluation is both mandatory and challenging.

Insight into patient satisfaction levels may serve as a catalyst for healthcare practitioners to deliver services that are more respectful and patient-centered^1,2^. Therefore, determining factors affecting patient-perceived satisfaction is critical for planning improvements in the health system^2^.

### 4.4. Satisfaction domains

Previous studies examining patient satisfaction, have determined that the components that increase it are measurable and tangible: assurance, reliability, responsiveness, and empathy, among others^1,2^. In this context, we propose that perceived satisfaction in the field of Fetal Therapy could be summarized in six domains: Follow-up care, environment, transparency, accessibility of care, link between physician and patient and global satisfaction.

These domains address those critical aspects that impact patients-perceived ’satisfaction. Each domain has also a number of items whose can be independently interpreted and assessed.

#### Domain 1: “Follow-up care”

It seeks to highlight the continuity of care by the same physician team throughout the care journey. We analyzed it through three items: the frequency the same professional attended the patient in the successive medical visits (question 1), the multidisciplinary management of the Unit through the participation of other specialists (pediatric surgeons, neonatologists…) (question 2), and the congruence and consistency of the information provided (question 3).

Parents who are faced with a fetal anomaly that is likely to decrease the length or quality of neonatal life should be extensively counseled by obstetrics and perinatal specialists^26^. Being attended by the same professional at the successive visits could influence the patient’s trust placed in the healthcare provider, valuing the Unit as a familiar and known place.

In the same line, Taillieu et al.^27^ reported that parents greatly appreciated a multidimensional support, both in the pre-to-postnatal period, independent of the outcome. Thus, we advocate for interventions to encourage healthcare staff to consider the participation of other specialists involved in the future management of the newborn, such as neonatologists, pediatric surgeons, pediatric cardiologists…

In our Unit, we have a specific clinic for the management of congenital heart disease. In that, the most complex cases are jointly assessed by a specific and clearly defined team comprising fetal medicine specialists, pediatric cardiologists, and pediatric surgeons, on a weekly basis. The same applies to the management of fetal gastrointestinal or respiratory anomalies.

This collaborative appraisal ensures the transmission of a clear, consistent, and shared information between the different professionals involved in patient care. It could also facilitate the informed shared decision-making processes, empowering parents from a sense of agency amidst uncertainties^27–29^.

#### Domain 2: “Environment”

Five questions helped us to study this domain. It encompassed the conditions of the Department’s facilities (question 4), the waiting time to be attended (question 5) the quiet, pleasant, and uninterrupted atmosphere of the consultation (question 6), the privacy and confidentiality during the medical visit (question 7) and the care given to the image by the Department’s personnel (question 8).

A comfortable and welcoming setting plays a pivotal role in shaping patient experiences^23^. Adedeji et al. reported that certain conditions, as cleanliness and maintenance of hygiene, are determinant of satisfaction^30^. Coincidentally, Kweon et al.^23^ suggested that the cleanliness of the rooms and the availability of quiet areas might directly impact patients ’comfort and their overall experience. Thus, investing in facilities that prioritize comfort and cleanliness could significantly impacts patients-perceived ’satisfaction^23^.

The same applies to waiting time at the clinic. Previous research carried out by Alqahtani et al. reported concerns regarding the longer waiting time patients experienced while seeking healthcare services^21^. Likewise, various patient satisfaction evaluation surveys have found waiting times a significant, ongoing source of dissatisfaction^21,24^. Patients expect to be seen by doctors on time as they are assigned a specific consultation time.

In fact, in our pilot study, waiting time is the only element that, according to the proposed scoring scale, was considered “satisfactory” (score 4.45). Accordingly, healthcare providers should reduce waiting time as much as possible.

Patient privacy is another criteria used to assess “environment” domain. It has been reported to demonstrate a strong positive correlation with patient satisfaction^21^. In our survey, the special care taken with privacy and confidentiality is perceived as a maximum strength, as 89.6% (26/29) of the surveyed gave a score of five. Therefore, respecting patient’s privacy is critical to a successful practice^21^.

In the same vein, we speculate that a quiet and pleasant atmosphere in the setting and care for the image by medical staff are two elements that also may have a significant influence in the patient perceptions of the hospital environment.

#### Domain 3: “Transparency”

In terms of providing information reliably and carefully, it is considered fundamental for a trustworthy physician-patient relationship. We suggested summarizing this domain into 3 items: the clarity of the explanation given regarding fetal conditions and concerns (question 9), the use of means to facilitate the understanding of the information such as drawings and explanatory diagrams (question 10) and the frequency with which professional asked questions to verify the information transmitted (question 11).

Transparency and clarity in patient education have been underscored as essential components, particularly in obstetrics research. A comprehensive patient education during prenatal care correlates with reduced anxiety and maternal stress, and enhanced self-efficacy^10^.

Moreover, patients expect to be fully informed of the diagnostic suspicion and all relevant information connected with it^31^. Counseling quality can have a significant effect on emotional and psychological outcomes specially in patients faced with difficult medical decisions^32^. Hence, a clear and accessible information empowers patients and facilitates their active participation in their healthcare journey^23^, leading to increase patient-perceived satisfaction.

A study conducted by Taillieu et al.^27^ analyzed patient-reported outcomes for parents with a lived experience of a prenatal diagnosis of isolated congenital diaphragmatic hernia. They reported that parents expressed a strong need for understanding of the condition in general and the treatment options. Some techniques as drawings and explanatory diagrams could draw the eye and entice patient engagement, as used in our program.

In alignment with our research, Crombag et al.^6^ conducted a study to explore parental feelings when considering maternal-fetal surgery. It reveals that some parents perceived that their concerns were not taken seriously by clinical staff.

Generally, patients expect to be treated well by a committed healthcare professional who takes their needs seriously and has a respectful approach^31^. Thus, it is therefore common practice in our Unit to ask questions and leave space for doubts and concerns. A communication technique usually used in clinic is, once the explanation has been completed, ask the patient to re-explain what she has understood. This feedback method helps the professional to ensure that the information has been properly grasped or, on the contrary, it should be adapted to the patient’s educational or intellectual level.

#### Domain 4: “Accessibility of care”

We consider that patients-perceived ‘satisfaction begins with the first contact with the Department. Therefore, we evaluated this domain through the following elements: the adequacy in time and form of being referred to the Fetal Medicine and Therapy Department (question 12), the willingness to provide a new appointment in case of a problem with the assigned date or time (question 13) and the suitability of the signage available to reach the Unit (question 14).

Correct information at a local level and good communication between local providers and the specialized center are an integral part of the care accesibility^27,31^. Crombag et al.^6^ informed that parents perceived a break in the continuity of care between basic and referral center caused additional stress. It highlights the importance of a smooth transition and clear and direct communication between centers.

Additionally, patients value prompt access to medical assistance, characterized by short to reasonable waiting times and accessibility through various channels^31^. To address this, our Department provides patients with a card containing contact information at their initial medical visit, ensuring assistance is readily available at any time.

It is noteworthy that in a referral center with several specialized subunits, patients value very positively the presence of signage that helps them to find their way around and get to their appointment properly. In fact, it was the most highly valued item in this domain.

#### Domain 5: “Link between physician and patient”

Communication skills are key in complex and transcendent decision-making situations. In this context, the emotional sphere of the professional gains great significance. Specifically, by the “link between physician and patient” domain we aimed to define the comprehensive and empathetic care to the patient, with special emphasis on emotional and psychological attention.

The difficulty in its analysis, led us to inquire through four questions that explore the kindness and courtesy of the professionals (question 15), the honesty, trust and security conveyed (question 16), the interest in understanding emotions and needs (question 17) and the accompaniment and support received by the medical team (question 18).

The attitude of medical staff has demonstrated a significant determinant of satisfaction^30^. Being treated with dignity, respect, and courtesy influence patients’ satisfaction with care^31^.

Interestingly, empathy skills also have implications in patients-perceived ’satisfaction. The ability to understand patients ’needs and emotions is crucial in situations of high emotional impact such as after the diagnosis of fetal anomaly. In fact, in our survey, the understanding of patients ’needs and knowledge of emotions, scored a mean of 4.34, revealing an only “satisfactory” rating.

It should be strengthened by undergoing training courses, acquiring, and improving communication skills. Pay more attention to psychological needs of patients may result in more empathy on their part^2^, underlining the vital role of healthcare providers as leading player in patients ’satisfaction.

Moreover, patient-perceived satisfaction has been also linked with the presence of a supportive and compassionate staff^33^. Compassion is now identified as an important, but previously, hidden ingredient of quality care, associated with patients’ overall ratings^34^. Improving it is an essential and potent means for enhancing care quality^34^. Consequently, we have prioritized the assessment of accompaniment and support received by the medical team within our analysis framework.

#### Domain 6: “Global satisfaction”

It aspired to reflect the general patient ‘evaluation of the entire care experience with Fetal Therapy program via two elements: The general satisfaction with the Program (question 19) and the willingness to recommend it (question 20).

### 4.5. Barriers for satisfaction analysis

Despite the importance of measuring satisfaction to improve the quality of care, the usefulness of perceived satisfaction surveys is debated, given the frequent sample biases and the lack of continuity in the tools used to reproduce them^35,36^.

Added to this, there is a conceptual problem. The term “satisfaction” could have diverse meanings for each patient, depending on their own level of expectation^35^.

Nevertheless, surveys are the most objective available method to evaluate care quality^35^. Through the results, physicians have informative feedback to modify those aspects poorly perceived by patients.

### 4.6. Clinical and research implications

The development of a tool to evaluate patient-perceived satisfaction is essential in any healthcare system that seeks continuous improvement. While general patient satisfaction instruments provide valuable insights, they may not capture the unique aspects of Fetal Therapy, such as the emotional and ethical challenges faced by patients.

Compared to existing tools, *“FETAL Surgery Global Satisfaction Tool*” offers a tailored approach to the experiences of patients undergoing Fetal Therapy. It may help to identify areas of weakness and implement targeted strategies.

Although initially tested in a single center and for a specific procedure (EXIT), it has the potential to be applicable to many other procedures performed in our Program (laser fetoscopy, intrauterine transfusion, myelomeningocele repair…), as well as, in other Fetal Medicine and Therapy Units worldwide that regularly perform fetal procedures.

It could facilitate the exchange of information and the standardization of a method for benchmarking, being the ultimate objective an improvement in care quality.

### 4.7. Strengths and limitations

This study provides a first in-depth exploration of maternal perceived satisfaction with a Fetal Medicine and Therapy program using a novel tool. One strength is the high participation of women (29/34) in the period under study. A further strong point is that women were involved in the design of the questionnaire, which strengthens the understanding and acceptance of the domains to the patients.

However, several limitations warrant acknowledgment. It was conducted in a single center and at a single period, so it is not possible to generalize the obtained results. Additionally, the present study findings might be interpreted with caution owing to the sampling approach, as pilot study was carried out with its limitations. To enhance its external validity, further research with larger and more diverse sample groups is essential.

In addition, the findings are based on Spanish population. It is uncertain whether the survey in unidimensional in other geographical areas. Thus, verification of cross-cultural validity will be required in the future.

This study is also limited in its recall period. There was a certain difference in the time elapsed between the healthcare experienced and the completion of the survey, and this could bias the results obtained according to their greater or lesser recall of the process. However, given that fetal procedures are applied for serious medical conditions with potential lifelong consequences and involve a high emotional impact, the influence on the results could be minimized. Moreover, the care program of the Department has not changed since the implementation of the Fetal Medicine and Therapy Program in 2007, which reinforces the results obtained.

Eventually, specific variables such as neonatal and maternal outcomes could impact survey answers. Nevertheless, we explored it and no statistically significant differences between participants and nonrespondents were observed.

## 5. Conclusions

Quality of care emerges as a paramount concern within any Fetal Medicine and Therapy Program committed to ongoing medical advancement. However, a validated tool for appraising care quality of patients undergoing Fetal Therapy is absent.

Thus, we propose “*FETAL Surgery Global Satisfaction Tool*” as a valuable instrument for assessing patient-perceived satisfaction. From a multidimensional approach, it encompasses input from six key domains. This tool may represent a step forward for Fetal Units capturing patients ’satisfaction as a key barometer of the quality of care and identifying areas for improvement.

Further studies are warranted to evaluate its validity and its applicability in different settings and patient populations.

## Data Availability

The datasets generated and analyzed during the current study are available from the corresponding author on reasonable request.

## Declarations

### Ethics approval and consent to participate

This study has been performed in accordance with the tenets of the Declaration of Helsinki and was approved by the Quality Department of Virgen del Rocio University Hospital, given written institutional permission. The institutional ethics committee (Andalusian Ethics Committee) confirmed that formal authorization on its part was not necessary. Informed consent was obtained from all the patients for clinical studies.

### Consent for publication

Not applicable.

### Authoŕs contributions

M.D., A.C., J.D., L.G., and G.A. contribute to conception and design. M.D. was responsible to acquisition of data. M.D., A.C., J.D., L.G., and G.A. contributed to the analysis and interpretation of data. All authors contribute to drafting the article or revising it critically for important intellectual content. All authors read and approved the final manuscript.

## Acknowledgements

All persons that contributed to this study are listed authors and meet the criteria for authorship.

## List of abbreviations

CREDES: Conducting and Reporting Delphi Studies
EXIT Ex: Utero Intrapartum Treatment
NHS: National Health Service

